# The SMART Text2FHIR Pipeline

**DOI:** 10.1101/2023.03.21.23287499

**Authors:** Timothy A. Miller, Andrew J. McMurry, James Jones, Daniel Gottlieb, Kenneth D. Mandl

## Abstract

**Objective:** To implement an open source, free, and easily deployable high throughput natural language processing module to extract concepts from clinician notes and map them to Fast Healthcare Interoperability Resources (FHIR).

**Materials and Methods:** Using a popular open-source NLP tool (Apache cTAKES), we create FHIR resources that use modifier extensions to represent negation and NLP sourcing, and another extension to represent provenance of extracted concepts.

**Results:** The SMART Text2FHIR Pipeline is an open-source tool, released through standard package managers, and publicly available container images that implement the mappings, enabling ready conversion of clinical text to FHIR.

**Discussion:** With the increased data liquidity because of new interoperability regulations, NLP processes that can output FHIR can enable a common language for transporting structured and unstructured data. This framework can be valuable for critical public health or clinical research use cases.

**Conclusion:** Future work should include mapping more categories of NLP-extracted information into FHIR resources and mappings from additional open-source NLP tools.

## Introduction and background

In the United States, billions of notes written by clinicians become a part of the electronic health record (EHR) each year. Notes contain a large percentage of the useful information in EHRs, when subjected to natural language processing (NLP).(1–7) While open source tools specialized to clinical NLP are in widespread use,(8–15) the variability in hospital data systems meant that each new NLP implementation has traditionally required substantial customization.

However, advances in both technology and regulation now make access to and processing of EHR-based text both turnkey and scalable. FHIR (Fast Healthcare Interoperability Resources) has become a de facto standard for presentation of data from EHR systems to external applications.

Further, interoperability provisions in the 21st Century Cures Act Rule require EHR vendors to support the SMART on FHIR(16,17) and SMART/HL7 Bulk FHIR Access (18) application programming interfaces (APIs). These interfaces allow standardized access to clinical text, along with structured information specified in the US Core for Data Interoperability (USCDI),(19) including laboratory testing, diagnoses, and medications.

This new data liquidity creates an unprecedented opportunity to apply NLP methods, which can convert unstructured text into structured information, for example, extracting from doctor’s notes, references to diagnoses, signs and symptoms, procedures, and the relations among these elements.

## Objective

We sought to implement an open source, free, high throughput, and easily deployable NLP module to extract concepts from clinician notes and map them to FHIR format. The result would be text-based information intermingled and used alongside structured information, allowing downstream users (e.g., clinical researchers, quality analysts, regulators, etc.) to manipulate this data source for a variety of use cases.

## Materials and Methods

Figure 1 shows an overview of the “SMART Text2FHIR” system. The core engine for “SMART Text2FHIR” is Apache cTAKES (clinical Text Analysis and Knowledge Extraction System),(8) an open-source clinical NLP software library for extracting information from unstructured text in EHRs. cTAKES is a highly configurable package with dozens of pipeline components that can be arranged by library users.

**Figure 1:**
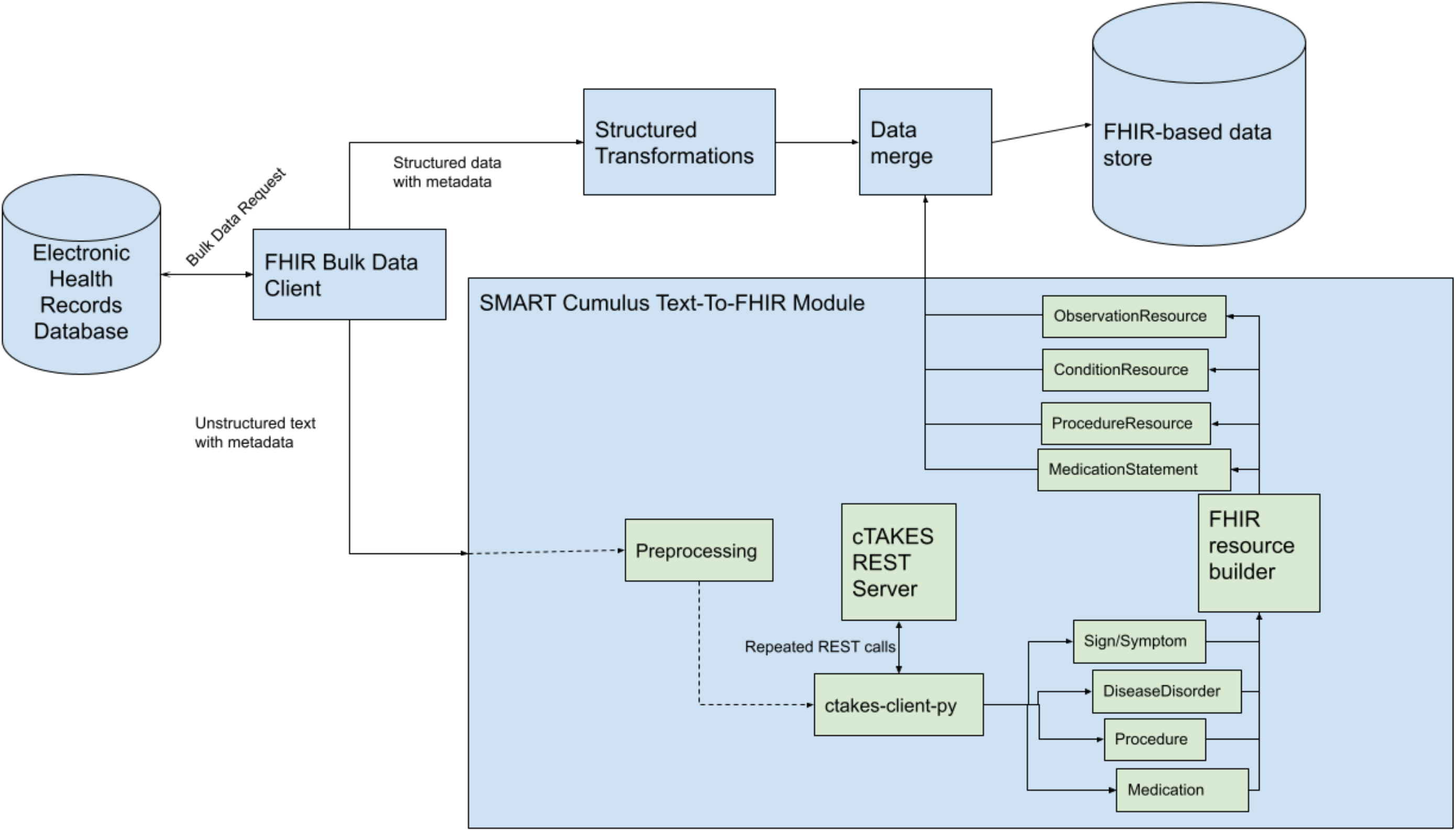
A diagram of the information flow that the SMART Text2FHIR module sits within.

Perhaps the most important cTAKES component is the dictionary lookup module, which maps strings in input texts to normalized codes in source terminologies, as well as Concept Unique Identifiers (CUIs) in the Unified Medical Language System (UMLS) for interoperability. The default cTAKES pipeline uses SNOMED-CT and RxNORM source dictionaries, and we make use of that pipeline for this work. We use a fast rule-based model, the cTAKES Context(20) implementation, for negation detection. This module labels each mapped concept with a *polarity* attribute, which is one by default and minus one if the concept is negated in the text (e.g., *no fever*).

This library uses standard FHIR resource types and annotates them with NLP-specific data and metadata. Extensibility is a core feature of FHIR; specifically, extensions provide a standard way for users to address specific use cases by adding elements to the core resource structures. Every FHIR resource and data element can be extended, and each extension contains a URI-based identifier and additional data (which may be comprised of other, nested extensions). Some FHIR elements can also be extended with “modifier” extensions, which alter the interpretation of the resource or element, and may not be ignored by applications or users of the data. SMART Text2FHIR uses both FHIR regular and modifier *extensions*.

Mappings were developed that convert outputs from broad semantic groups in the UMLS (and the cTAKES “type system” which closely mirrors it) to FHIR resources. Procedures are mapped to the Procedure Resource, Signs/Symptoms are mapped to Observation Resources, Diseases/Disorders are mapped to Condition Resources, and Medications are mapped to MedicationStatement Resources.

Next, extensions to track provenance of NLP outputs were implemented – one important attribute of our implementation is that the automatic and less-certain nature of NLP-extracted resources is highlighted, and that provenance information is preserved so that downstream users can manually validate or even replicate NLP system behavior if required for their use cases.

We track the NLP system version used to produce the FHIR resource with an “nlp-source” modifier extension that includes “algorithm” and “version” values. The algorithm field is a human-readable description of the algorithm, and the version field is a URL pointing to the location of the NLP software. Representing this information as a modifier extension rather than a regular extension is important, as this helps indicate to downstream users that these concepts (e.g., codes representing diseases) do not have the same epistemological status as mappings from coded data, and need to be treated differently by any systems that ingest them.

To track the position of each extracted NLP concept in its original document, we added a *derivation-reference* extension which has been proposed for inclusion in the next release of FHIR; it has integer-typed *offset* and *length* fields to represent character offsets of the concept in the source document, which, in combination with the *reference* field that points to the original document, would allow a downstream user to find the original text span corresponding to the extracted concept.

Finally, a modifier extension to represent negation of concepts called “nlp-polarity” was also implemented. Negation is common in EHR text, as it is often important to, for example, rule out a competing diagnosis or explicitly assert the absence of a symptom. This was implemented as a modifier extension since it fundamentally changes the meaning of the concept and contains a boolean value withthe value *False* for negated concepts and *True* otherwise.

## Results

SMART Text2FHIR is an extension to Apache cTAKES that performs NLP-to-FHIR mappings with easily obtainable open-source artifacts. cTAKES has an existing REST server implementation, which returns a json-formatted output string with an ad hoc data model. Our new modifications include creating a GitHub repository for building a Docker image of the cTAKES REST server, a publicly available pre-built Docker image from that repository, and an easily installable python client that implements the mapping from the cTAKES REST output string to our proposed FHIR representations. We use these components as follows:

### Publicly available Docker image

This publicly available Docker image^1^ is built from the ctakes-covid-container repository on Github.^2^ This sets up the cTAKES REST API with a customized NLP pipeline enriched to be able to find COVID-19-related signs and symptoms. The relevant components of this pipeline are a dictionary lookup that finds mentions of terms in SNOMED-CT, RxNORM, and a customized COVID-19 dictionary and maps them to UMLS CUIs (for SNOMED-CT and RxNORM) or custom codes. The cTAKES pre-built dictionaries require UMLS authentication at container start-up, so this image accepts a UMLS API key as an environment variable in its run command.

### cTAKES REST client

Once the Docker container running the cTAKES REST server is started, it can be called by a python library that we have developed called ctakes-client-py.^3^ This client library can be installed with the pip (package installer for python) tool. The client provides functions to simplify REST calls to the server and organizes the returned JSON data by semantic type. The text2fhir module of the client provides methods for converting each semantic type into the appropriate resource. This module also provides a single simplified entry point that takes the unstructured FHIR in and returns a collection of FHIR resources that are ready for downstream use.

### Example outputs

Figure 2 shows a snippet of an example input text, and the resulting FHIR output. The full example can be seen in the online supplement.

**Figure 2:**
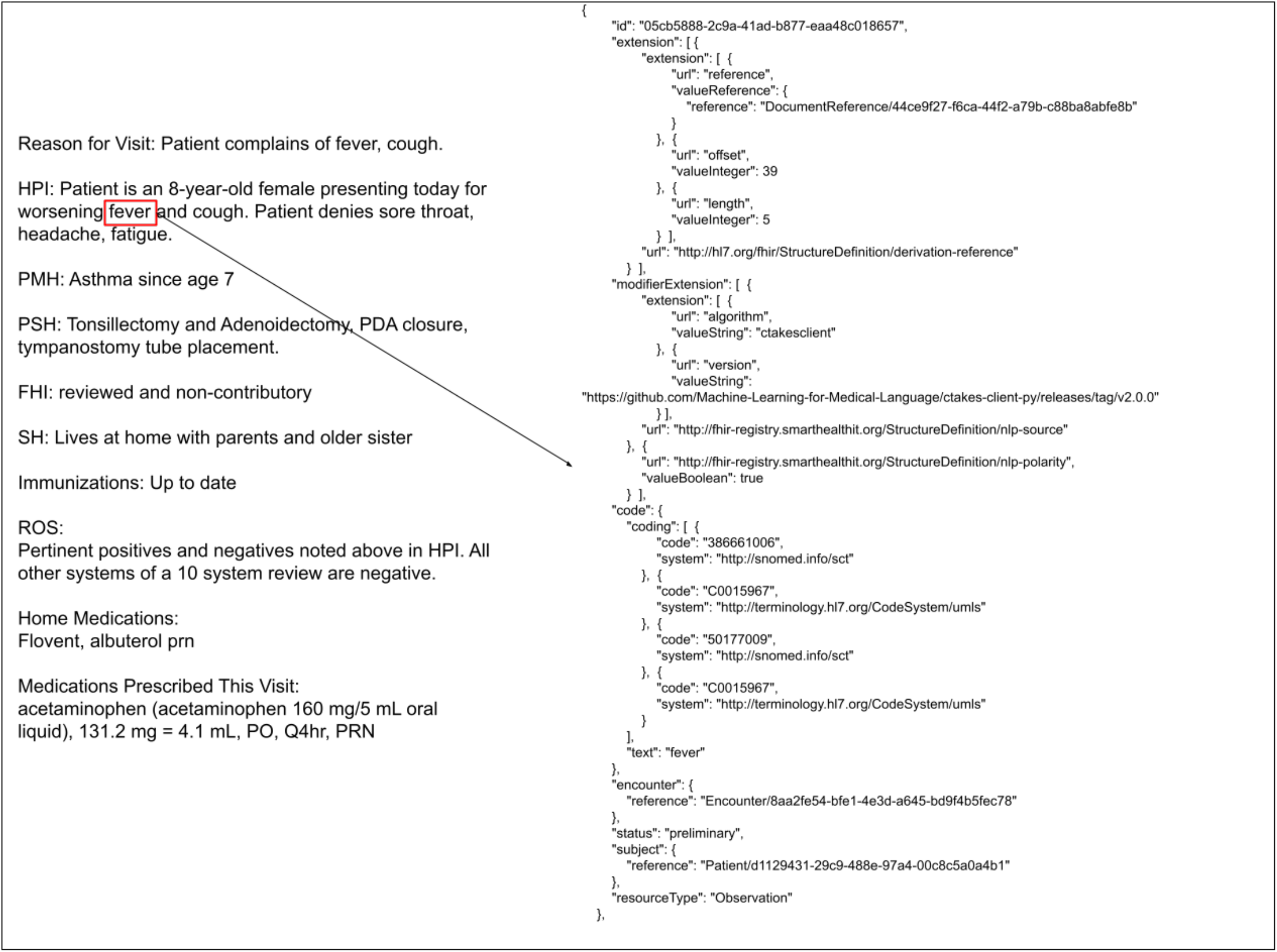
An example snippet of a note (left), with multiple clinical concepts. The concept “fever” is converted to a FHIR resource represented by the JSON text on the right.

## Discussion

We anticipate that SMART Text2FHIR can address many use cases that require aligning clinical text data alongside structured data. For example, in a multisite network, multiple collaborating research sites could run the same NLP to FHIR process, either pooling their data in a shared HIPAA-compliant cloud instance or as separate on-premises stores. The NLP output from all sites would then be in the same format from the start. This could, for example, serve as a foundation for sharing classifiers that operate over clinical text (21) at multiple centers. Another example use case is outside researchers who want to run de-identified queries against EHR data (e.g., regulators or public health officials): however they formulate the query, the mapping to the internal FHIR representation can be re-used or shared across sites.

There are a few known open-source attempts to map from NLP outputs to FHIR. Apache cTAKES has a built-in ctakes-fhir module that can convert many elements of the cTAKES data model into FHIR resources. However, that implementation makes use only of the Basic Resource type, which offers great flexibility in adapting to new data types, but is more general than might be desired for many data types that already have more appropriate mappings to FHIR resources. This will make it difficult for users to leverage existing FHIR knowledge when reviewing NLP derived data, as well as add complexity when working with structured and unstructured data in a single study.

The NLP2FHIR system(22) contains several logical mappings from multiple NLP systems into FHIR resources, including mapping disease/disorder mappings to ConditionResource, mapping signs and symptoms to ObservationResource, and mapping procedures to ProcedureResource. However, one major missing element of this effort was a lack of provenance data – that is, the ability to work backwards from FHIR resources to the original source to validate NLP outputs if necessary. It also requires manual installation of several packages it depends on, making setup somewhat tricky. Our solution includes package-managed software installs that remove the difficulty of manually managing software versions in dependent libraries.

There are also existing commercial offerings from Microsoft (23) and Amazon Web Services (24), but our goal is to produce an open source alternative that can run economically at scale and be maintained by the research and FHIR communities.

## Conclusion

The open-source software implementation is available on GitHub. We encourage readers to join the community developing these mappings to ensure future mappings are done in a logical and transparent manner.

Currently, negation (i.e., polarity) is the only status represented, but there are NLP methods for detecting, for example, uncertain/hedged hypothetical, and non-patient-related concepts that could easily be integrated using similar mechanisms.

While we focused on one widely used clinical NLP tool, cTAKES, extending to other tools requires minimal effort: First, a Docker wrapper that sets up a REST endpoint (we have previously implemented such a library for Medspacy^4^), and then an output mapping step. This could take the form of a conversion into the cTAKES ad hoc data model as a postprocess in the REST server, or the forking of cTAKES server-specific functions in the ctakes-client-py library.

Future work must include mapping more categories of NLP-extracted information into FHIR resources, for example, temporal information. Clinical notes often contain narrative information describing the disease course, with mentions of past and future (potential) events, and representing these important temporal details is crucial for many use cases.

## Data Availability

This manuscript does not describe created data but software resources, which are available at open source repositories linked in the manuscript.

## Funding acknowledgement

Work reported in this publication was supported by three contracts 90AX0022, 90AX0019, 90AX0031, and 90C30007 from the Office of the National Coordinator of Health Information Technology; the Centers for Disease Control and Prevention of the U.S. Department of Health and Human Services (HHS) as part of a financial assistance award (The contents are those of the author(s) and do not necessarily represent the official views of, nor an endorsement, by CDC/HHS, or the U.S. Government). A cooperative agreement from the National Center for Advancing Translational Sciences U01TR002623; Grants from the National Library of Medicine (R01LM012973, R01LM012918); and the Boston Children’s Hospital PrecisionLink Initiative. The content is solely the responsibility of the authors and does not necessarily represent the official views of the funding sources.

## Footnotes

1 https://hub.docker.com/r/smartonfhir/ctakes-covid

2 https://github.com/Machine-Learning-for-Medical-Language/ctakes-covid-container

3 https://github.com/Machine-Learning-for-Medical-Language/ctakes-client-py

4 https://github.com/Machine-Learning-for-Medical-Language/medspacy-covid-container

